# Early Detection of Cognitive Decline in Parkinson’s Disease Using Natural Language Processing of Clinical Notes: A Systematic Review and Meta-Analysis Protocol

**DOI:** 10.1101/2025.10.11.25337798

**Authors:** Ravi Shankar, Ziyu Goh, Xu Qian

**Author notes:** **Corresponding Author:** Dr Ravi Shankar; Clinical Research & Innovation Office, Tan Tock Seng Hospital, National Healthcare Group, Singapore, 308433, Email correspondence.

## Abstract

**Background:** Cognitive decline affects approximately 40% of Parkinson’s disease (PD) patients within 10 years of diagnosis, progressing to dementia in 80% of patients after 20 years. Early detection of cognitive changes is crucial for timely intervention and care planning. Clinical notes contain rich, longitudinal information about cognitive symptoms that may precede formal diagnosis, yet this unstructured data remains largely unexplored for systematic cognitive decline detection.

**Objective:** To systematically review and meta-analyze studies using natural language processing (NLP) techniques to detect early cognitive decline in Parkinson’s disease patients from clinical notes, evaluating the accuracy, timing, and predictive features of different NLP approaches.

**Methods:** This systematic review will follow PRISMA guidelines and search PubMed, Embase, Web of Science, IEEE Xplore, ACL Anthology, and PsycINFO from inception to December 2025. Eligible studies must apply NLP methods to clinical notes for detecting or predicting cognitive decline in PD patients. Two reviewers will independently screen studies using Covidence software. Quality assessment will use PROBAST+AI for prediction model studies and QUADAS-2 for diagnostic accuracy studies. Meta-analysis will pool diagnostic accuracy measures using bivariate random-effects models where appropriate.

**Data Synthesis:** Narrative synthesis will describe NLP methodologies, feature extraction techniques, and validation approaches. Meta-analysis will calculate pooled sensitivity, specificity, and area under the curve (AUC) for studies with comparable outcomes. Subgroup analyses will examine performance by NLP approach (rule-based vs. machine learning vs. deep learning), cognitive domain, and time to diagnosis.

**Discussion:** This review will establish the current evidence base for NLP-based early detection of cognitive decline in PD, identify optimal methodological approaches, and guide future development of clinical decision support tools for cognitive monitoring in PD populations.

## Introduction

Parkinson’s disease represents one of the most prevalent neurodegenerative disorders globally, affecting approximately 1% of individuals over 60 years and characterized by both motor and non-motor symptoms that progressively impair quality of life [1]. While traditionally conceptualized as a movement disorder, cognitive impairment has emerged as one of the most significant non-motor features, with mild cognitive impairment (PD-MCI) present in 25-30% of patients at diagnosis and progressing to Parkinson’s disease dementia (PDD) in up to 80% of patients within 20 years [2]. The cognitive domains most commonly affected include executive function, attention, visuospatial abilities, and memory, with the pattern and progression varying considerably among patients [3]. Early detection of cognitive decline in PD is critical for clinical management, as it influences medication choices, enables timely psychosocial interventions, facilitates care planning, and provides opportunities for clinical trial enrollment when neuroprotective strategies may be most effective [4].

The current approach to detecting cognitive decline in PD relies primarily on periodic neuropsychological assessments, typically administered annually or when cognitive concerns arise, using standardized batteries such as the Montreal Cognitive Assessment (MoCA) or comprehensive neuropsychological testing according to Movement Disorder Society criteria [5]. However, this approach has significant limitations including the resource-intensive nature of comprehensive testing, practice effects with repeated assessments, limited sensitivity to subtle early changes, and the episodic nature of formal assessments that may miss gradual decline between visits [6]. Furthermore, access to neuropsychological expertise varies considerably across healthcare settings, creating disparities in cognitive monitoring for PD patients [7].

Clinical documentation represents an underutilized resource for cognitive monitoring in PD, as healthcare providers routinely document observations about patients’ cognitive status, functional abilities, and daily living challenges in clinical notes during regular visits [8]. These notes contain rich longitudinal data capturing subtle cognitive changes reported by patients, caregivers, and clinicians, often predating formal cognitive diagnosis by months or years [9]. The narrative descriptions in clinical notes may capture early cognitive symptoms such as word-finding difficulties, organizational problems, or medication management issues that do not yet meet thresholds for formal cognitive impairment diagnosis but signal emerging decline [10]. The increasing adoption of electronic health records (EHRs) has created vast repositories of digital clinical text that can be analyzed systematically using computational methods to identify patterns indicative of cognitive decline [11].

Natural language processing has emerged as a powerful approach for extracting meaningful information from unstructured clinical text, with applications ranging from phenotyping to outcome prediction across various medical conditions [12]. In the context of neurodegenerative diseases, NLP techniques have shown promise in identifying cognitive symptoms from clinical notes in Alzheimer’s disease and related dementias, achieving high accuracy in detecting documented cognitive impairment and predicting future cognitive decline [13]. The application of NLP to PD-related cognitive decline presents unique opportunities and challenges, as the cognitive profile in PD differs from other dementias, often featuring prominent executive dysfunction and fluctuating attention that may manifest differently in clinical documentation [14].

Recent advances in NLP methodology, particularly the development of transformer-based models and clinical language models, have substantially improved the ability to understand context and extract complex medical concepts from clinical text [15]. These methods can identify not only explicit mentions of cognitive symptoms but also subtle linguistic patterns, temporal relationships, and contextual information that may indicate emerging cognitive decline [16]. Studies have begun exploring these approaches in PD populations [17-20], but the evidence remains fragmented across different methodologies, populations, and outcome definitions, necessitating systematic synthesis to understand the current state of knowledge and identify optimal approaches.

The integration of NLP-based cognitive monitoring into clinical practice could transform early detection and management of cognitive decline in PD by providing continuous, automated screening of clinical documentation, identifying patients at risk for cognitive decline before obvious symptoms emerge, reducing the burden on healthcare systems while improving detection rates, and enabling personalized risk stratification and targeted intervention strategies [21]. However, successful implementation requires understanding the accuracy, reliability, and generalizability of different NLP approaches, as well as their integration potential within existing clinical workflows [22-24].

### Objectives

The primary objective of this systematic review and meta-analysis is to comprehensively evaluate the diagnostic accuracy and predictive performance of natural language processing techniques applied to clinical notes for early detection of cognitive decline in Parkinson’s disease patients, synthesizing evidence across different NLP methodologies, clinical settings, and patient populations to determine the overall effectiveness and optimal approaches for clinical implementation.

Specific objectives include: (1) identifying and categorizing NLP methods used for cognitive decline detection in PD, including rule-based systems, traditional machine learning, and deep learning approaches, with assessment of their relative performance characteristics; (2) evaluating the diagnostic accuracy metrics including sensitivity, specificity, positive predictive value, negative predictive value, and area under the receiver operating characteristic curve for different NLP approaches in detecting both mild cognitive impairment and dementia in PD populations; (3) examining the temporal aspects of NLP-based detection, specifically how early these methods can identify cognitive decline relative to clinical diagnosis and whether certain linguistic features or patterns provide earlier warning signs; (4) identifying the specific linguistic features, semantic patterns, and contextual information most predictive of cognitive decline in PD to understand the mechanisms through which NLP systems achieve detection; and (5) assessing methodological quality, risk of bias, and generalizability of existing studies to provide evidence-based recommendations for future research and clinical application development.

## Methods

### Study Design

This systematic review and meta-analysis will adhere to the Preferred Reporting Items for Systematic Reviews and Meta-Analyses (PRISMA) guidelines and the Meta-analysis of Observational Studies in Epidemiology (MOOSE) recommendations to ensure methodological rigor and transparent reporting [25, 26]. The protocol will be registered with PROSPERO prior to commencing the review to establish a priori methods and minimize risk of selective reporting bias. Given the interdisciplinary nature of NLP applications in healthcare, the review will incorporate guidance from both medical informatics and clinical epidemiology perspectives to comprehensively evaluate technical performance and clinical validity.

The review will employ a two-stage approach, first conducting a systematic review to identify and qualitatively synthesize all relevant studies, followed by meta-analysis of studies with sufficient homogeneity in methods and outcomes. This approach recognizes the methodological diversity in NLP research while enabling quantitative synthesis where appropriate to generate pooled estimates of diagnostic accuracy and predictive performance [27].

### Search Strategy

A comprehensive search strategy will be developed in collaboration with a medical librarian experienced in both biomedical and computer science databases. The search will span multiple databases to capture publications from medical, informatics, and computational linguistics communities. Primary databases will include PubMed/MEDLINE, Embase, Web of Science Core Collection, IEEE Xplore Digital Library, ACM Digital Library, and the ACL Anthology. Additional searches will target PsycINFO for neuropsychological perspectives and Google Scholar for grey literature including conference proceedings and preprints.

The search strategy will combine three conceptual domains using Boolean operators: (1) Parkinson’s disease terms including “Parkinson’s disease,” “Parkinson disease,” “Parkinsonism,” “PD,” and “Parkinsonian disorders”; (2) cognitive decline concepts including “cognitive impairment,” “cognitive decline,” “dementia,” “cognitive dysfunction,” “mild cognitive impairment,” “MCI,” “PDD,” “neuropsychological,” and related terms; and (3) natural language processing methodology including “natural language processing,” “NLP,” “text mining,” “clinical notes,” “electronic health records,” “EHR,” “machine learning,” “deep learning,” “transformers,” “BERT,” and “clinical documentation.” No date or language restrictions will be applied initially, though non-English publications will be assessed for translation feasibility. The search will cover publications from database inception through December 2025.

Grey literature searches will include ProQuest Dissertations & Theses Global, conference proceedings from relevant meetings (AMIA, EMNLP, ACL, MDS Congress), and preprint servers (medRxiv, bioRxiv, arXiv). Forward and backward citation searching will be conducted for all included studies and relevant systematic reviews. Authors of conference abstracts and ongoing studies will be contacted to identify completed but unpublished work.

### Eligibility Criteria

Studies will be included if they meet all of the following criteria: (1) Population - patients with diagnosed Parkinson’s disease, including idiopathic PD, with or without cognitive impairment at baseline; studies including mixed populations must report PD-specific results separately; (2) Intervention/Exposure - application of any NLP technique to analyze clinical notes, including physician notes, nursing documentation, or multidisciplinary team notes from electronic health records or medical charts; studies may use rule-based, machine learning, or hybrid approaches; (3) Comparator - studies must include a reference standard for cognitive status, which may include formal neuropsychological testing, clinical diagnosis of PD-MCI or PDD according to established criteria, validated cognitive screening tools, or expert clinical judgment documented in the medical record; (4) Outcomes - studies must report diagnostic accuracy metrics (sensitivity, specificity, AUC) or predictive performance measures for detecting or predicting cognitive decline, mild cognitive impairment, or dementia in PD patients; (5) Study Design - observational studies including cohort, case-control, and cross-sectional designs; both retrospective and prospective studies will be included.

Studies will be excluded if they: analyze only structured EHR data without clinical text; focus solely on speech or writing samples rather than clinical documentation; examine cognitive symptoms in other parkinsonian disorders without separate PD analysis; use NLP only for cohort identification without cognitive outcome assessment; are case reports, editorials, or opinion pieces without empirical data; or lack sufficient information to extract diagnostic accuracy measures. Conference abstracts will be included only if sufficient methodological detail is available or can be obtained from authors.

### Study Selection

Study selection will be managed using Covidence systematic review software to ensure standardized screening and transparent documentation of the selection process. After removing duplicates, two reviewers will independently screen titles and abstracts against the eligibility criteria using a standardized screening form. The form will be piloted on 50 randomly selected citations and refined to ensure consistent application. Disagreements will be resolved through discussion, with a third reviewer consulted when consensus cannot be reached.

Full-text screening will follow the same dual-review process, with reviewers documenting specific reasons for exclusion according to the predefined criteria. For studies with unclear eligibility, authors will be contacted for clarification with a two-week response window. Non-English studies meeting initial criteria will undergo assessment for translation resources before final inclusion decisions. A PRISMA flow diagram will document the number of studies at each selection stage with reasons for exclusion clearly specified.

### Data Extraction

A standardized data extraction form will be developed and piloted on five studies representing different NLP approaches to ensure comprehensive capture of relevant information. The form will be implemented in Covidence to facilitate systematic extraction and comparison across studies. Two reviewers will independently extract data from all included studies, with discrepancies resolved through discussion or third-party arbitration.

Extracted data will include: study characteristics (authors, year, country, setting, study design, sample size, data sources, time period); population characteristics (PD diagnostic criteria, disease duration, baseline cognitive status, age, sex distribution, medications); NLP methodology (approach type, specific algorithms, feature extraction methods, preprocessing steps, software/tools used, validation approach); cognitive outcomes (definition of cognitive decline, diagnostic criteria used, assessment methods, time to diagnosis); performance metrics (sensitivity, specificity, PPV, NPV, AUC, accuracy, calibration measures); temporal aspects (prediction horizon, time from documentation to diagnosis, longitudinal vs. cross-sectional analysis); and implementation details (computational requirements, external validation, code availability, integration with clinical systems).

For studies reporting multiple models or outcomes, data will be extracted for all reported analyses to enable subgroup comparisons. When studies report performance at multiple thresholds, the threshold optimizing sensitivity and specificity balance will be selected as the primary outcome, with sensitivity analyses examining alternative thresholds.

### Quality Assessment

Methodological quality and risk of bias will be assessed using tools appropriate to the study design and research question. The Prediction model Risk Of Bias ASsessment Tool for Artificial Intelligence (PROBAST+AI) will be used for studies developing or validating prediction models, evaluating risk of bias across four domains: participants, predictors, outcome, and analysis [28]. The Quality Assessment of Diagnostic Accuracy Studies-2 (QUADAS-2) tool will be applied to diagnostic accuracy studies, assessing patient selection, index test, reference standard, and flow/timing [29].

Additional quality considerations specific to NLP studies will be evaluated using adapted criteria from the Transparent Reporting of a multivariable prediction model for Individual Prognosis Or Diagnosis for Artificial Intelligence (TRIPOD+AI) statement, including: data preprocessing transparency, feature engineering documentation, model selection justification, handling of class imbalance, cross-validation approach, and external validation [30]. Studies will also be assessed for potential biases specific to NLP applications including selection bias from EHR data availability, information bias from documentation quality variations, and temporal bias from changes in documentation practices over time.

Two reviewers will independently conduct quality assessments, with disagreements resolved through consensus discussion. Quality assessment results will not determine study inclusion but will inform sensitivity analyses and GRADE assessment of evidence certainty. A summary table will present quality assessment results across all included studies, identifying patterns of methodological strengths and limitations.

### Data Synthesis and Analysis

Data synthesis will begin with a narrative summary describing the characteristics of included studies, NLP methodologies employed, and cognitive outcomes assessed. Studies will be grouped by NLP approach (rule-based, traditional machine learning, deep learning, hybrid) and cognitive outcome (MCI detection, dementia detection, cognitive decline prediction) to identify patterns and methodological trends. Forest plots will visualize individual study results even when meta-analysis is not appropriate, providing a comprehensive overview of the evidence landscape.

Meta-analysis will be conducted when at least three studies report comparable outcomes using similar NLP approaches and reference standards. Diagnostic accuracy meta-analysis will employ hierarchical bivariate random-effects models to jointly synthesize sensitivity and specificity, accounting for correlation between these measures and threshold effects [31]. Summary receiver operating characteristic (SROC) curves will be constructed to visualize overall diagnostic performance. For predictive models reporting time-to-event outcomes, hazard ratios will be pooled using generic inverse-variance methods.

Heterogeneity will be assessed using the I^2^ statistic and tau-squared, with values above 50% indicating substantial heterogeneity warranting investigation. Planned subgroup analyses will examine: NLP approach (rule-based vs. machine learning vs. deep learning), cognitive domain (executive vs. memory vs. global), prediction horizon (<1 year vs. 1-3 years vs. >3 years), clinical setting (specialized movement disorder clinics vs. general neurology vs. primary care), and documentation type (physician notes vs. multidisciplinary notes). Meta-regression will explore associations between study characteristics and performance metrics when sufficient studies are available.

Sensitivity analyses will examine the impact of: excluding studies at high risk of bias, alternative meta-analysis models (fixed-effects, Bayesian approaches), different thresholds for defining cognitive decline, and excluding studies without external validation. Publication bias will be assessed using funnel plots and Egger’s test when at least 10 studies are included in meta-analysis. The certainty of evidence will be evaluated using the GRADE approach, considering risk of bias, inconsistency, indirectness, imprecision, and publication bias.

## Discussion

This systematic review and meta-analysis will provide the first comprehensive synthesis of evidence on using natural language processing of clinical notes for early detection of cognitive decline in Parkinson’s disease, addressing a critical gap in understanding how computational approaches can enhance cognitive monitoring in this vulnerable population. The findings will have important implications for clinical practice, research methodology, and health technology development, potentially transforming approaches to cognitive surveillance in PD through automated analysis of routine clinical documentation.

The clinical significance of this review extends beyond academic interest to practical applications in PD management, where early detection of cognitive decline could substantially impact patient outcomes through timely intervention, medication optimization, and care planning support. Current clinical practice relies heavily on episodic formal cognitive assessments that may miss gradual changes between visits, creating opportunities for NLP-based continuous monitoring to identify at-risk patients earlier in the disease trajectory [32]. The synthesis of diagnostic accuracy metrics across studies will establish benchmarks for NLP performance, enabling clinicians to understand the reliability and limitations of these tools compared to traditional assessment methods. Understanding which linguistic features and documentation patterns are most predictive of cognitive decline will also enhance clinical awareness of early warning signs that may be documented but not fully recognized in routine practice.

The methodological insights generated by this review will advance the field of clinical NLP by identifying best practices for applying these techniques to neurodegenerative cognitive syndromes, where the gradual onset and heterogeneous presentation create unique challenges compared to other medical conditions. By systematically comparing different NLP approaches from rule-based systems to state-of-the-art transformer models, the review will elucidate the trade-offs between interpretability and performance, computational requirements and accuracy, and generalizability versus specificity that influence implementation decisions [33]. The analysis of temporal aspects will be particularly valuable, revealing how far in advance NLP methods can detect cognitive decline and whether certain approaches are better suited for near-term versus long-term prediction, critical considerations for clinical utility and intervention planning.

Several challenges and limitations are anticipated in synthesizing this emerging literature, reflecting both the novelty of applying NLP to PD cognitive outcomes and the inherent complexity of analyzing clinical documentation. The heterogeneity in NLP methodologies, ranging from simple keyword searches to complex deep learning models, may limit quantitative synthesis and necessitate careful subgroup analyses to generate meaningful conclusions. Variations in reference standards for cognitive impairment, from brief screening tools to comprehensive neuropsychological batteries, introduce additional complexity in comparing diagnostic accuracy across studies [34]. The quality and completeness of clinical documentation likely varies across healthcare settings and providers, potentially affecting NLP performance in ways that are difficult to quantify or adjust for in meta-analysis.

The review findings will have important implications for future research directions in both clinical NLP and PD cognitive assessment. Expected identification of evidence gaps will highlight priority areas for methodological development, such as handling longitudinal documentation, incorporating multimodal data, or developing PD-specific language models that better capture the unique cognitive phenotype of this condition [35]. The synthesis may reveal needs for standardization in evaluation metrics, reference standards, and reporting practices to enable better comparison and reproduction of NLP studies in neurodegenerative diseases. Understanding current limitations will also inform development of hybrid approaches that combine NLP insights with other clinical data sources for more comprehensive cognitive risk assessment.

Implementation considerations emerging from this review will be crucial for translating research findings into clinical practice, as technical performance alone does not guarantee successful integration into healthcare workflows. The review will examine not only accuracy metrics but also practical factors such as computational requirements, need for local adaptation, and integration with existing EHR systems that influence feasibility of deployment [36]. Identifying studies that have progressed from development to implementation will provide valuable lessons about barriers and facilitators to clinical adoption, informing strategies for future translation efforts. The review will also consider equity implications, as NLP performance may vary across patient subgroups based on factors such as education, language proficiency, or healthcare access that influence documentation quality.

The potential impact of NLP-based cognitive monitoring on healthcare delivery models for PD warrants careful consideration, as these tools could enable new paradigms of proactive cognitive care rather than reactive diagnosis and management. Automated screening of clinical notes could identify patients requiring formal cognitive assessment more efficiently than current referral patterns, potentially reducing disparities in access to specialized neuropsychological services [37]. The ability to track cognitive trajectories through continuous analysis of routine documentation could support personalized medicine approaches, tailoring interventions based on individual patterns of decline rather than population averages. Integration with clinical decision support systems could alert providers to concerning cognitive changes and prompt evidence-based interventions at optimal timepoints.

Ethical and regulatory considerations surrounding the use of NLP for cognitive assessment in PD will require careful attention as these technologies move toward clinical implementation. Issues of algorithmic bias, particularly if models are trained on data from limited populations, could perpetuate or exacerbate healthcare disparities if not carefully addressed through diverse training data and rigorous validation across patient subgroups [38]. Privacy concerns related to automated analysis of clinical notes, especially for sensitive cognitive information, necessitate robust data governance frameworks and transparent communication with patients about how their data is being used. Regulatory pathways for NLP-based cognitive assessment tools remain evolving, with questions about whether these constitute medical devices requiring formal approval and how to ensure ongoing performance monitoring post-deployment.

This systematic review will provide essential evidence synthesis to guide the responsible development and implementation of NLP tools for cognitive monitoring in Parkinson’s disease, balancing innovation potential with rigorous evaluation of accuracy, reliability, and clinical utility. By comprehensively examining the current state of evidence, identifying knowledge gaps, and highlighting methodological considerations, the review will accelerate progress toward precision medicine approaches for cognitive care in PD while ensuring that technological advances translate into meaningful benefits for patients and caregivers facing the challenges of cognitive decline in neurodegenerative disease.

## Data Availability

All data produced in the present work are contained in the manuscript

## References

1. Dorsey, E.R., et al., The Emerging Evidence of the Parkinson Pandemic. J Parkinsons Dis, 2018. 8(1): p. S3–s8.

2. Aarsland, D., et al., Parkinson disease-associated cognitive impairment. Nature Reviews Disease Primers, 2021. 7(1): p. 47.

3. Goldman, J.G. and E. Sieg, Cognitive Impairment and Dementia in Parkinson Disease. Clin Geriatr Med, 2020. 36(2): p. 365–377.

4. Weintraub, D., et al., The neuropsychiatry of Parkinson’s disease: advances and challenges. Lancet Neurol, 2022. 21(1): p. 89–102.

5. Litvan, I., et al., Diagnostic criteria for mild cognitive impairment in Parkinson’s disease: Movement Disorder Society Task Force guidelines. Mov Disord, 2012. 27(3): p. 349–56.

6. Skorvanek, M., et al., Global scales for cognitive screening in Parkinson’s disease: Critique and recommendations. Mov Disord, 2018. 33(2): p. 208–218.

7. Willis, A.W., et al., Neurologist care in Parkinson disease: a utilization, outcomes, and survival study. Neurology, 2011. 77(9): p. 851–7.

8. Ford, E., et al., Extracting information from the text of electronic medical records to improve case detection: a systematic review. J Am Med Inform Assoc, 2016. 23(5): p. 1007–15.

9. Locke, S., et al., Natural Language Processing in Medicine: A Review. Trends in Anaesthesia and Critical Care, 2021. 38.

10. Chandler, C., et al., An explainable machine learning model of cognitive decline derived from speech. Alzheimers Dement (Amst), 2023. 15(4): p. e12516.

11. Wu, S., et al., Deep learning in clinical natural language processing: a methodical review. J Am Med Inform Assoc, 2020. 27(3): p. 457–470.

12. Kreimeyer, K., et al., Natural language processing systems for capturing and standardizing unstructured clinical information: A systematic review. J Biomed Inform, 2017. 73: p. 14–29.

13. Amra, S., et al., Derivation and validation of the automated search algorithms to identify cognitive impairment and dementia in electronic health records. J Crit Care, 2017. 37: p. 202–205.

14. Zhan, A., et al., Using Smartphones and Machine Learning to Quantify Parkinson Disease Severity: The Mobile Parkinson Disease Score. JAMA Neurol, 2018. 75(7): p. 876–880.

15. Elvas, L.B., A. Almeida, and J.C. Ferreira, Natural language processing in medical text processing: A scoping literature review. International Journal of Medical Informatics, 2025. 204: p. 106049.

16. Li, Y., et al., Clinical-Longformer and Clinical-BigBird: Transformers for long clinical sequences. 2022.

17. Aresta, S., et al., Digital phenotyping of Parkinson’s disease via natural language processing. npj Parkinson’s Disease, 2025. 11(1): p. 182.

18. Twala, B., AI-driven precision diagnosis and treatment in Parkinson’s disease: a comprehensive review and experimental analysis. Front Aging Neurosci, 2025. 17: p. 1638340.

19. Hossain, M.A., E. Traini, and F. Amenta Machine Learning Applications for Diagnosing Parkinson’s Disease via Speech, Language, and Voice Changes: A Systematic Review. Inventions, 2025. 10, DOI: 10.3390/inventions10040048.

20. Rabie, H. and M.A. Akhloufi, A review of machine learning and deep learning for Parkinson’s disease detection. Discover Artificial Intelligence, 2025. 5(1): p. 24.

21. Dahodwala, N., et al., Treatment disparities in Parkinson’s disease. Ann Neurol, 2009. 66(2): p. 142–5.

22. Velupillai, S., et al., Using clinical Natural Language Processing for health outcomes research: Overview and actionable suggestions for future advances. J Biomed Inform, 2018. 88: p. 11–19.

23. Shankar, R., A. Bundele, and A. Mukhopadhyay, A Systematic Review of Natural Language Processing Techniques for Early Detection of Cognitive Impairment. Mayo Clinic Proceedings: Digital Health, 2025. 3(2): p. 100205.

24. Shankar, R., A. Bundele, and A. Mukhopadhyay, Natural language processing of electronic health records for early detection of cognitive decline: a systematic review. npj Digital Medicine, 2025. 8(1): p. 133.

25. Page, M.J., et al., The PRISMA 2020 statement: an updated guideline for reporting systematic reviews. Bmj, 2021. 372: p. n71.

26. Stroup, D.F., et al., Meta-analysis of observational studies in epidemiology: a proposal for reporting. Meta-analysis Of Observational Studies in Epidemiology (MOOSE) group. Jama, 2000. 283(15): p. 2008–12.

27. Debray, T.P., et al., A guide to systematic review and meta-analysis of prediction model performance. Bmj, 2017. 356: p. i6460.

28. Moons, K.G.M., et al., PROBAST+AI: an updated quality, risk of bias, and applicability assessment tool for prediction models using regression or artificial intelligence methods. BMJ, 2025. 388: p. e082505.

29. Whiting, P.F., et al., QUADAS-2: a revised tool for the quality assessment of diagnostic accuracy studies. Ann Intern Med, 2011. 155(8): p. 529–36.

30. Collins, G.S., et al., TRIPOD+AI statement: updated guidance for reporting clinical prediction models that use regression or machine learning methods. BMJ, 2024. 385: p. e078378.

31. Reitsma, J.B., et al., Bivariate analysis of sensitivity and specificity produces informative summary measures in diagnostic reviews. J Clin Epidemiol, 2005. 58(10): p. 982–90.

32. Leroi, I., et al., Cognitive impairment in Parkinson disease: impact on quality of life, disability, and caregiver burden. J Geriatr Psychiatry Neurol, 2012. 25(4): p. 208–14.

33. Wang, Y., et al., Clinical information extraction applications: A literature review. J Biomed Inform, 2018. 77: p. 34–49.

34. Dubois, B., et al., Diagnostic procedures for Parkinson’s disease dementia: recommendations from the movement disorder society task force. Mov Disord, 2007. 22(16): p. 2314–24.

35. Gonzalez-Latapi, P. and C. Marras, Epidemiological Evidence for an Immune Component of Parkinson’s Disease. J Parkinsons Dis, 2022. 12(1): p. S29–s43.

36. Sheikhalishahi, S., et al., Natural Language Processing of Clinical Notes on Chronic Diseases: Systematic Review. JMIR Med Inform, 2019. 7(2): p. e12239.

37. Mantri, S., et al., State-level prevalence, health service use, and spending vary widely among Medicare beneficiaries with Parkinson disease. npj Parkinson’s Disease, 2019. 5(1): p. 1.

38. Chen, I.Y., et al., Ethical Machine Learning in Healthcare. Annu Rev Biomed Data Sci, 2021. 4: p. 123–144.

